# IL-17A, IFN-γ, and MIP-3α Plasma Profiles Predict Clinical Stage Transition in First-Episode Psychosis

**DOI:** 10.64898/2026.02.17.26346145

**Authors:** Miguel Rosado, Catarina Empadinhas, Vitor Santos, Cátia Santa, Mário Grãos, Manuel Coroa, Sofia Morais, Miguel Bajouco, Helder Costa, Inês Baldeiras, Artur Paiva, Antonio Macedo, Nuno Madeira, Bruno Manadas

## Abstract

**Background:** Early detection of individuals at risk for clinical deterioration in first-episode psychosis (FEP) remains a vital challenge in psychiatric care. Emerging evidence indicates that immune dysregulation might play a crucial role in the pathophysiology and progression of psychotic disorders.

**Aims:** This study examined the predictive potential of a plasma cytokine and chemokine panel in anticipating clinical stage transition of FEP patients.

**Method:** Using multiplex immunoassays, plasma samples from a cohort of 35 FEP patients were screened for the quantification of 21 analytes. Participants were clinically assessed at baseline and follow-up and classified according to a validated staging model. Data was used to predict clinical stability over a 12-month follow-up period.

**Results:** IL-17A was found to be significantly increased in transitioning patients (p = 0.045), with a medium standardized effect size and wide confidence interval (Hedges’ g = - 0.687, 95% CI [-1.379, 0.004]). A logistic regression model was determined, which revealed that higher baseline levels of IL-17A were significantly linked to progression to a more advanced clinical stage, while higher baseline levels of MIP-3α and IFN-γ were associated with clinical stability. This combined cytokine model exhibited strong predictive capacity (AUC = 0.853), indicating its potential as a biomarker panel for early risk assessment.

**Conclusions:** These findings highlight the importance of neuroimmune mechanisms in the development of psychotic disorders and advocate for the inclusion of immunological markers within staging-based models of care. Incorporating cytokine profiling into clinical practice could improve personalised treatment strategies and lead to better long-term outcomes for individuals with FEP.

## INTRODUCTION

### Psychotic Disorders and the Need for Early Diagnostic Precision

Psychotic disorders are serious mental illnesses that rank among the leading causes of disability on a global scale [1]. Studies have shown that individuals with psychosis exhibit greater functional impairments, significantly higher suicide rates [2], and an increased risk of premature death compared to the general population [3]. Although these conditions have a significant impact, most research has predominantly focused on schizophrenia (SCZ), leaving other major psychotic disorders relatively understudied [4]. Primary psychotic disorders include SCZ, schizoaffective disorder, schizotypal disorder and delusional disorder, among others. These disorders are defined by impairments affecting one or more of the following key functional domains: delusions, hallucinations, disorganised thinking (speech), abnormal motor behaviour (including catatonia), and negative symptoms. Each disorder is defined by specific diagnostic criteria that consider factors such as illness duration and symptom presentation heterogeneity. Nonetheless, delineating clear nosological boundaries remains challenging, thereby complicating differential diagnosis and hindering the development of targeted treatment strategies, for which accurate classification is essential [5, 6]. First-episode psychosis (FEP) represents a critical focus of research, as improved understanding and early diagnostic precision have the potential to substantially enhance long-term outcomes through the implementation of timely and targeted interventions. Despite the implementation of numerous clinical and research programs focused on FEP as a nosological entity, subsequent diagnostic evolution is often heterogeneous, and there is still no consensus on its definition [7]. The underlying etiologies can be categorised into primary and secondary causes. Primary causes include the consequent development of mental illnesses such as SCZ, schizoaffective disorder and mood disorders, while secondary causes encompass conditions like substance-induced psychosis and psychotic disorders due to another medical condition [8]. A clinically and functionally oriented staging model has been proposed for psychotic disorders [9, 10], incorporating stages from 0 to 4. Stage 0 represents the presence of genetic risk factors without symptoms, aligning with the premorbid phase in the clinical staging model. Stage 1 includes individuals considered to be at high clinical risk for developing psychosis (CHR-P), characterised by early warning signs or subtle symptoms, while stage 2 is characterised by isolated occurrences of FEP in those who achieve early and full recovery. Stage 3 is associated with a progressive trajectory characterised by delayed or incomplete recovery. Stage 4, on the other hand, represents a severe, persistent, and unremitting illness corresponding to the chronic phase in this model. Although interventions exist for each stage of the disease, their application is often challenged by significant diagnostic and prognostic uncertainties. This highlights the importance of identifying molecular biomarkers that can predict disease progression, enabling earlier diagnosis and providing more precise guidance for therapeutic options, particularly pharmacological interventions. One promising biomarker is the blood cytokine profile, which may provide valuable insights into disease progression and support treatment planning.

### Inflammation and Cytokines in Psychosis

In recent years, there has been a significant increase in the number of publications regarding the potential role of inflammatory states in the development of mental disorders [11]. Studies have consistently found elevated levels of pro-inflammatory cytokines in patients with SCZ, both at the onset and later stages of the illness. Interestingly, these abnormalities vary between the acute and stable phases, with specific cytokines showing potential as biomarkers for disease progression, while others indicate specific clinical features [12]. This phenomenon is associated with the function of the blood-brain barrier (BBB), which acts as a boundary between the central nervous system (CNS) and peripheral circulation, where inflammatory mediators and immune cells flow [13]. Cytokines can cross into the CNS under physiological conditions, such as during pregnancy [14], as well as during pathological states characterised by peripheral inflammation that compromises BBB integrity [13]. Among the various markers under consideration, elevated IL-17 levels in the blood of individuals with FEP have been reported [15, 16]. This pro-inflammatory cytokine, primarily produced by CD4+ T helper 17 (Th17) cells, intraepithelial lymphoid γδ T cells (γδ IELs), mucosal innate lymphoid cells type 3 (ILC3) and, to a lesser extent, neutrophils, plays a crucial role in infection control and mucosal defence [17–20]. IL-17-producing cells can mature in the thymus, bone marrow, or peripheral tissues and are distributed in the blood and various peripheral tissues, particularly in the gut. Indeed, gut dysbiosis and increased intestinal permeability are thought to activate these immune cells, triggering the release of IL-17 into the systemic circulation and potentially affecting brain function through cytokine signalling and disruption of the blood-brain barrier [21]. Additionally, stress, genetic predisposition, and environmental factors may further amplify IL-17 production, highlighting a complex interplay between immune activation, gut-brain communication, and neuroinflammation in the SCZ pathophysiology [22, 23].

### IL 17 and Other Cytokines as Potential Biomarkers

It is essential to investigate whether IL-17 elevation in SCZ is associated with clinical stage transition, as this could provide valuable insights into disease progression and identify new avenues for therapeutic interventions aimed at maintaining immune balance and preventing chronic inflammation [24]. Previous studies suggest that additional cytokines may contribute to the pathophysiology of drug-naïve, first-episode SCZ, including IFN-γ, IL-6, and IL-2 [16, 25]. Some studies have shown that the plasmatic profile of interleukins, such as an increase of IL-6 concentration, may be associated with poor treatment response and the presence or absence of negative symptoms [26]. Despite growing research into the role of cytokines in the onset of FEP, the existing literature predominantly emphasises comparison between patient cohorts and healthy controls [25, 27, 28]. There are still significant inconsistencies about which specific cytokines best predict disease progression. This variability highlights the urgent need for further investigation to clarify the precise roles of cytokines in psychosis, especially to understand how these markers affect the transition from early psychosis to more chronic stages of the disease. To address these issues, this study compared the plasma cytokine profiles of patients with FEP who have been prospectively followed for at least one year, aiming to identify specific analytes associated with disease stage transition that could serve as predictive biomarkers. By identifying these inflammatory mediators, we aimed to provide critical insights into earlier and more targeted interventions, facilitating personalised treatment strategies and a better understanding of the disease’s biological mechanisms. These findings have the potential not only to advance knowledge of FEP pathogenesis but also to elucidate the role of the immune system in psychotic disorders, supporting the development and contributing to more effective strategies for managing disease progression.

## MATERIALS AND METHODS

### Patient Selection

Blood samples were obtained from a cohort of individuals with first-episode psychosis (FEP) after written informed consent. The authors assert that all procedures contributing to this work comply with the ethical standards of the relevant national and institutional committees on human experimentation and with the Helsinki Declaration of 1975, as revised in 2013. All procedures involving human subjects/patients were approved by the Ethics Committee of the Faculty of Medicine, University of Coimbra (CE-122/2015). The inclusion criteria were: (a) individuals aged between 18 and 45 years; (b) ability to provide informed consent; (c) contact with a mental health department due to FEP (psychotic symptoms suggestive of a DSM-5 [5] diagnosis of SCZ, schizoaffective disorder, schizophreniform disorder, psychotic disorder non-otherwise specified, or brief psychotic disorder). The exclusion criteria were: (a) patients who had previous psychotic episodes; (b) individuals with substance-induced psychotic disorder; (c) neuropsychiatric symptoms arising from a CNS or other medical conditions.

The cohort was subsequently divided into two groups: patients with FEP who transitioned to stage 3 after at least 1 year of follow-up (group T), and patients with FEP who remained in stage 2 after at least 1 year of follow-up (group nT) [9, 10]. Sample IDs shown in this article are anonymized and were not known to anyone outside the research group.

### Cytokine Quantification

Plasma samples were analysed using an xMAP technology-based Milliplex Human High Sensitivity T Cell 21 Plex PREMIXED Magnetic Bead Kit (Merck - EMD Millipore cat. no. HSTCMAG28SPMX21), following the manufacturer’s instructions. The panel consisted of the following analytes: Fractalkine, GM-CSF, IFN-γ, IL-1β, IL-2, IL-4, IL-5, IL-6, IL-7, IL-8, IL-10, IL-12 (p70), IL-13, IL-17A, IL-21, IL-23, ITAC, MIP-1α, MIP-1β, MIP-3α and TNF-α. Readings were performed using a Bio-Plex 200 System and the software Bio-Plex Manager 5 (both from Bio-Rad). Post-acquisition data analyses were performed using Bio-Plex Manager 5, and all calibration curves were Five Parameter Logistic Curves (5PL). Standards, quality controls and blanks were run in duplicate, while samples were run in single wells.

### Data Analysis

Unless otherwise specified, all data manipulation, visualisation, and statistical analyses were performed in R (version 4.5.1) using the packages cited herein [29–46].

Sample groups were inspected for balance according to sex, drug usage and age with Fisher’s Exact Test for Count Data, Pearson’s Chi-squared test with Yates’ continuity and Wilcoxon-Mann-Whitney test with Benjamini-Hochberg p-value correction, respectively (Figure 1B). Normality of age distribution per group was evaluated before prior to the aforementioned test with Anderson–Darling test (Supplementary Table 1) and quantile–quantile plots (Supplementary Figure 1).

**Figure 1.**
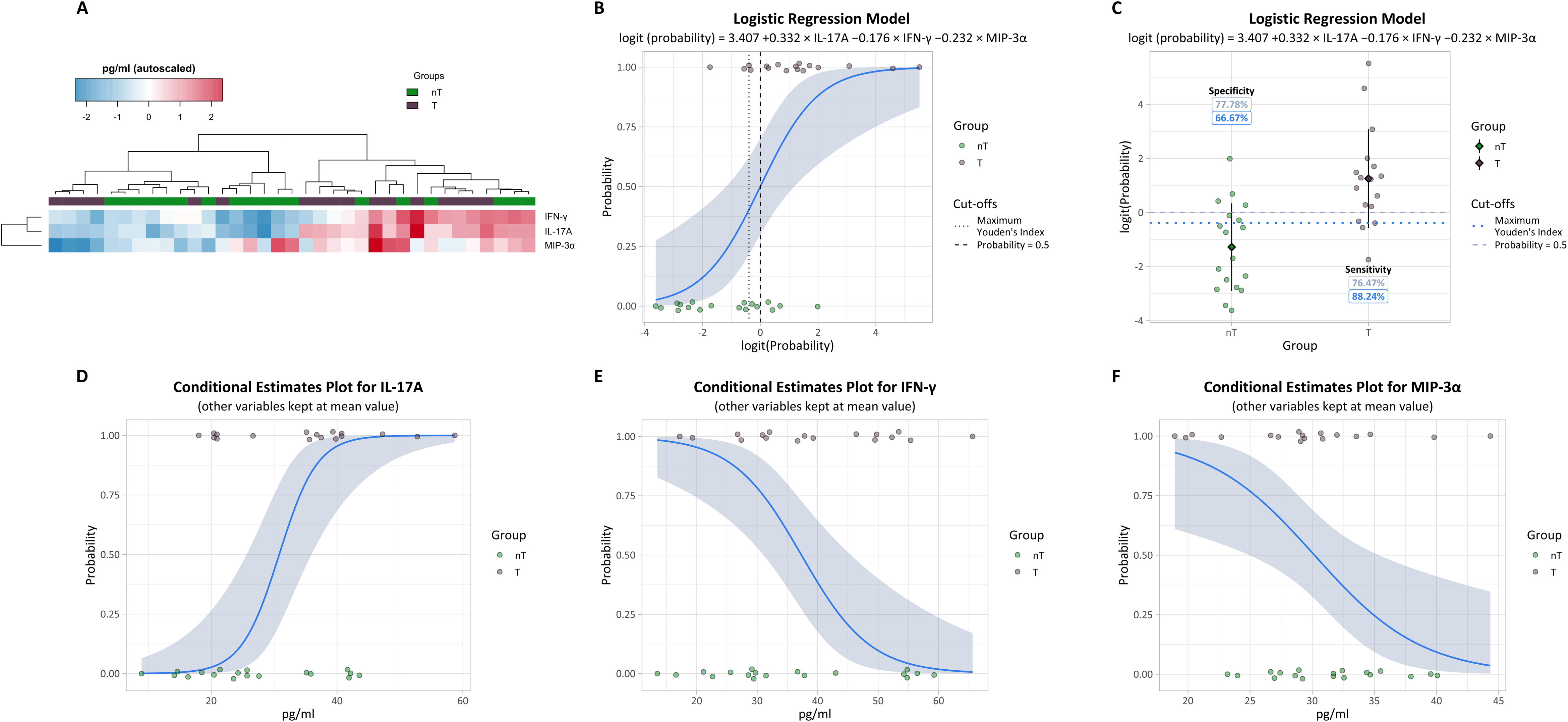
Description of the study population and univariate data analysis. (A) Layout of the sample selection process used in the study design. (B) Statistical analysis table of covariates in the study population. The symbols *, #, and $ indicate that p-values are derived from Fisher’s Exact Test for Count Data, Pearson’s Chi-squared test with Yates’ continuity correction, and Wilcoxon-Mann-Whitney test with Benjamini-Hochberg correction, respectively. (C) Box plot of IL-17A absolute quantification across the studied groups. The symbol * indicates a p-value below 0.05 according to Student’s t-Test. (D) Spearman’s correlation heatmap of samples using selected variables after data pre-processing. Values in each cell show Spearman’s rank correlation coefficient for each sample pair, rounded to two decimal places.

Feature selection was carried out in three sequential steps (Supplementary Figure 2): (i) removal of variables with missing or extrapolated values, (ii) exclusion of variables producing uncorrelated samples, and (iii) selection of statistically relevant variables using stepwise Linear Discriminant Analysis (sLDA), implemented in IBM® SPSS® Statistics (Version 22, Trial).

Exploratory analyses were conducted to identify potential outliers and problematic variables. Principal Component Analysis (PCA) was performed in MetaboAnalyst 6.0 [47] (Supplementary Figure 3), while Spearman’s rank correlations were calculated to inspect relationships between samples (Supplementary Figure 4, Supplementary Tables 2–5). Clustering analysis was performed on autoscaled data (mean-centred and variance-normalised) using Ward’s hierarchical agglomerative method [48] (Supplementary Figure 5).

Univariate statistical analysis was performed based on the data distribution per variable (Supplementary Table 6) to use the appropriate statistical test (Supplementary Tables 7 and 8). To prepare for multivariate modelling, data distributions (Supplementary Tables 9-11) were further assessed, and variables were log10-transformed where appropriate.

Multivariate modelling was performed using the features selected by sLDA to construct a multivariate logistic regression model to discriminate between experimental groups. Model performance and validity were evaluated through several complementary approaches: (i) variable coefficients were assessed using Wald tests, and multicollinearity was inspected with the Variance Inflation Factor and Tolerance (Supplementary Table 12); (ii) overall goodness of fit was determined by the likelihood ratio test and Hosmer–Lemeshow test (Supplementary Table 13); (iii) the assumption of linearity between predictors and the logit function was verified using the Box–Tidwell procedure and Component-plus-Residual plots (Supplementary Table 14 and Supplementary Figure 6).

Diagnostic performance was evaluated by Receiver Operating Characteristic (ROC) curve analysis, with the optimal classification threshold determined by the maximum Youden’s index.

Comparison of several machine learning pipelines (Supplementary Table 15) was performed using Orange Data Mining Software (version 3.37.0) [49].

## RESULTS AND DISCUSSION

### Plasma IL-17A is Altered in First Episode Psychosis with Clinical Stage Transition

The initial cohort comprised 56 individuals with FEP who underwent a baseline clinical assessment and blood sampling. For this analysis, only 35 patients were selected, as they were the only ones with more than one year of follow-up. These patients were then divided into two groups: FEP patients with no Clinical Stage Transition (n = 18, nT) and FEP patients with Clinical Stage Transition (n = 17, T) (Figure 1A). A detailed breakdown of the population by age, sex, and drug consumption is provided, showing no significant differences between the groups (Figure 1B).

The data processing pipeline (Supplementary Figure 2) began with the inspection of the raw quantification values (Supplementary Table 16) for each analyte in the assay, which revealed missing or extrapolated values for three analytes (IL-13, IL-8, and MIP-1α). Consequently, these analytes were excluded from further analysis. The remaining dataset was subjected to outlier analysis, revealing two uncorrelated samples (Supplementary Figures 3 and 4, and Supplementary Tables 2 and 3). Further examination of the overall cytokine profile (Supplementary Figure 5) revealed that four variables (IL-6, IL-1β, TNF-α, and IL-5) were noticeably increased exclusively in these samples, while the remaining analytes generally followed the same scale as in other samples. This indicated technical variability; therefore, these analytes were excluded from further analysis. These procedures retained all 35 observations for 14 analytes, yielding a highly correlated dataset (Figure 1D and Supplementary Tables 4 and 5) without sacrificing statistical power, as no samples were excluded. Univariate data analysis (Supplementary Tables 6, 7, and 8) revealed that only one cytokine, IL-17A, was significantly altered (p < 0.05) between T and nT plasma samples (Figure 1C).

### IL-17A, IFN-***γ*** and MIP-3***α*** May Predict Clinical Stage Transition in First Episode Psychosis

To further explore if any multi-analyte panel of potential biomarkers could be extracted from the cytokine profiling data, multivariate data analysis for feature selection, modelling, and sample classification was conducted. Stepwise Linear Discriminant Analysis (sLDA) demonstrated that three analytes, IL-17A, IFN-γ and MIP-3α, could effectively distinguish patients with FEP who transitioned to stage 3 (T) from those who remained in stage 2 (nT) (Supplementary Figure 7), with a sensitivity of 82.35% and specificity of 72.22%, indicating a significant diagnostic capability (Supplementary Figure 7B). Additionally, after cross-validation, 71.4% of all samples were correctly classified (Supplementary Table 17).

However, since several assumptions of sLDA weren’t met, the discriminant function was ignored for classification purposes, and this exploratory analysis was instead used to select features for the next step in the analytical pipeline. These assumptions included: (i) multivariate normality (Supplementary Table 9), which was not achieved even after data transformation (Supplementary Table 10), although this process did reduce the number of univariate non-normally distributed features in the dataset (Supplementary Tables 6 and 11); (ii) an inadequate sample-to-feature ratio; and (iii) equality of variance-covariance matrices (Supplementary Table 18), which, although met, depended on a very small sample size.

The overall pattern of these three cytokines across all samples (Figure 2A) suggested the distinction of two clusters: one with an overall relative decrease in these analytes, mainly consisting of nT samples, and another with an overall relative increase, mostly comprising T samples. A closer examination revealed that these clusters could be subdivided into subclusters, each with distinct cytokine abundance and sample composition profiles, suggesting that the relationship between clinical stage transitions and this cytokine panel may not be strictly linear.

**Figure 2.**
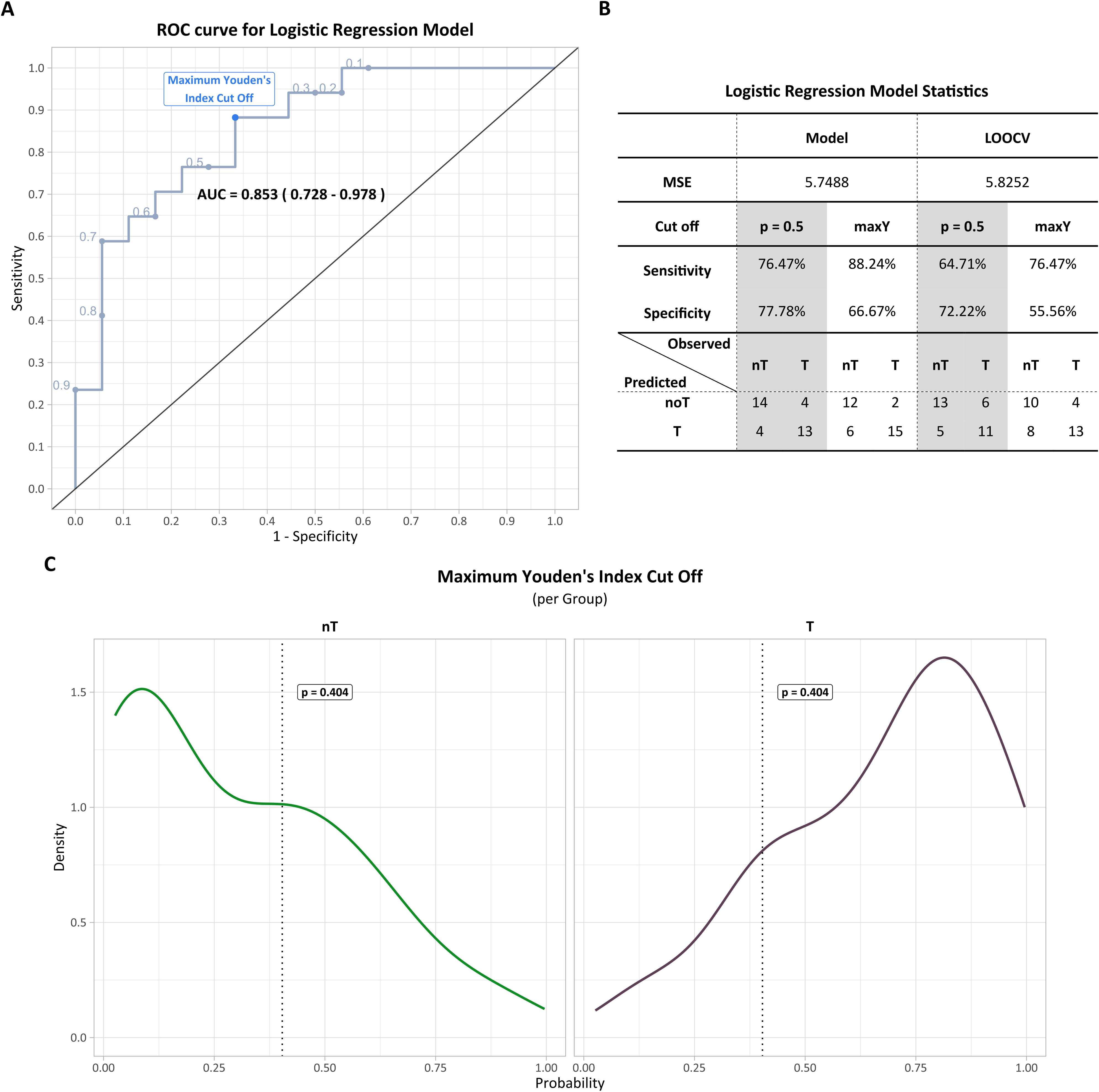
Selected discriminating features and logistic regression model visualisation. (A) Clustered heatmap of features selected for input into the logistic regression model with dendrograms. Intensity values are expressed as standardised and mean-centred (autoscaled) concentration values. Both samples and features were clustered according to Ward’s linkage method. (B) Sigmoid curve of the logistic regression model and dot plot of the study groups. The group “T” was considered as the positive class. The blue-transparent area indicates the 95% confidence interval of the model’s sigmoid curve. Vertical lines represent different probability cut-off limits used for classification. (C) Dot plot of the logistic regression model’s predictions for the logarithm of the odds ratio values across the study groups. Horizontal lines denote different probability cut-off limits used for classification. Lines and values in grey or blue refer to cut-offs and respective specificity and sensitivity values for 0.5 probability or maximum Youden’s J Index probability, respectively. (D), (E), and (F) show conditional estimates plots with grouped dot plots for IL-17A, IFN-γ, and MIP-3α, respectively. In each case, the sigmoid curve and respective 95% confidence interval (shown as a solid blue line and transparent blue area, respectively) are plotted according to the logistic regression model’s formula, using the respective feature’s values for all samples, while keeping the remaining features constant at their mean value.

To decide which analytical procedure to use next, the data were subjected to a series of machine learning pipelines, including both supervised and unsupervised methods, to identify the most reliable approach to cross-validation (Supplementary Figure 8, Supplementary Table 15). Assessment of each model’s diagnostic ability revealed that although logistic regression did not initially demonstrate the greatest diagnostic capacity, it proved to be the most resistant to cross-validation.

Thus, to better evaluate the relationship between the selected cytokines and the transition to the clinical stage in FEP, these analytes were incorporated into a logistic regression model (Figure 2B). Several assumptions and model validation checks were carried out (Supplementary Figure 6, Supplementary Tables 13 and 14). Although IL-17A and IFN-γ showed some degree of collinearity (Supplementary Table 12), the model was still used for sample classification. Using the conventional cut-off with a predicted probability of 0.5, the model achieved 77.78% specificity and 76.47% sensitivity, while the cut-off determined by the maximum Youden’s J Index (maxY) method yielded a specificity of 66.67% and a sensitivity of 88.24% (Figure 2C and 3B). When these cytokines were used for classification through a logistic regression model (Figure 3A), the diagnostic performance was comparable to that of the sLDA approach (Supplementary Figure 7).

**Figure 3.**
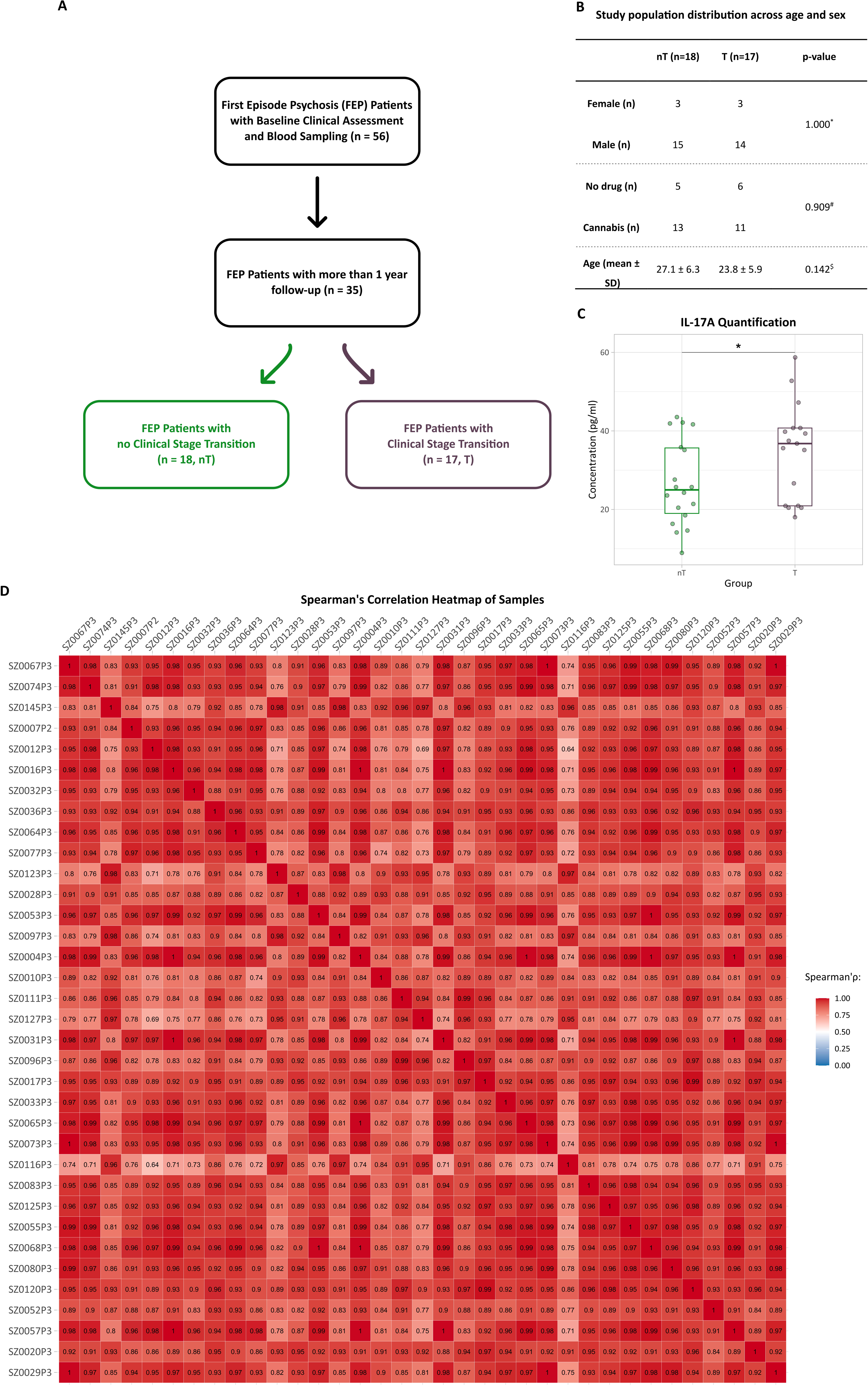
Evaluation of the logistic regression model’s diagnostic capacity. (A) Receiver operating characteristic curve of the logistic model. The area under the curve and the respective 95% confidence interval are shown in bold black text. The probability cut-off determined at the maximum Youden’s J Index is indicated by a blue dot. Other probability cut-offs are illustrated in grey along the curve. (B) Logistic regression model classification performance statistics, cross-validation, and confusion matrix. (C) Distribution of probability values predicted by the logistic regression model across the study groups. The dotted line and bold black text indicate the probability cut-off established at the maximum Youden’s J Index. LOOCV – leave-one-out cross-validation, MSE – mean squared error, p – probability, maxY – probability at maximum Youden’s J Index.

Furthermore, using the maxY method to define the cut-off (Figure 3C), rather than the conventional 0.5 threshold, resulted in higher sensitivity (88.24%) and yielded a classification model that was more robust to cross-validation (Figure 3B). However, this came at the cost of the overall value and robustness to cross-validation of specificity. It is evident that this cut-off is more effective for an approach prioritising sensitivity over specificity (Figure 3C).

Individual assessment of each analyte’s contribution to the model revealed two patterns: (i) a clear effect of IL-17A (Figure 2D), with higher values associated with T classification; and (ii) an unclear effect of IFN-γ and MIP-3α (Figure 2E and 2F), with higher values linked to nT classification. This suggests that the chosen threshold of 0.404 offers a better balance between sensitivity and specificity, optimising the model’s ability to distinguish between individuals who transitioned and those who did not, though it does increase the number of false positives.

### Prior Evidence Supports the Role of Cytokines in Clinical Stage Transition

Previous studies comparing healthy controls with patients experiencing FEP have emphasised the crucial role of inflammation and immune dysfunction in psychotic disorders. A systematic review reported increased levels of cytokines, such as IL-17 and IFN-γ, in patients with FEP compared to healthy controls [26]. However, other studies found no significant differences in IL-17 concentrations between the groups [24, 27]. Additionally, further studies have shown that patients with FEP display lower IL-17 levels compared to healthy controls [50, 51]. It was also reported that Th-17 immune response facilitates progression of inflammation in SCZ, associating IL-17 with cognitive impairment, and hypothesizing anti-IL-17A therapies as possible strategies for the treatment of endotoxemia-induced neuroinflammation and cognitive dysfunction [52].

To date, few studies have been conducted to compare groups of patients with FEP to assess the impact of cytokine concentrations on clinical stage transition. Among the limited studies comparing patients with FEP, one used a multiple logistic regression analysis with the Structured Interview for Prodromal Syndromes (SIPS) score, with IL-17 and TNF-β as independent variables. This study evaluated individuals (n=49) at ultra-high risk (UHR) for psychosis, corresponding to stage 1 of the clinical staging model. The results showed an increased Odds Ratio for both IL-17 and TNF-β, suggesting that higher levels of these cytokines were associated with a greater likelihood of clinical stage transition over a 2-year follow-up, compared to those who did not transition [53]. These findings are consistent with the results of our study. Remarkably, our study explores the transition from stage 2 to stage 3, capturing a more advanced phase of early psychosis. This distinction highlights its pioneering and innovative contribution, as it offers insight into a crucial yet understudied stage of disease progression.

Regarding the relationship between prolonged active psychosis and levels of pro-inflammatory cytokines, fewer studies have been conducted, and the available data indicate that an increase in MIP-3α serum levels is significantly associated with a longer initial time to remission, while a decrease in serum levels is related to a shorter time to remission [54]. Similarly, applying our model could provide valuable insights into the relationship between cytokine levels and treatment outcomes with antipsychotic medications. Specifically, testing the model may assist in identifying patients at higher risk of extended active psychosis, enabling more personalised therapeutic approaches and optimising treatment strategies during the early stages of psychosis.

## CONCLUSIONS

This study explored the predictive role of IL-17A, MIP-3α, and IFN-γ plasma levels in clinical stage transition among patients with FEP. Among the cytokines analysed, a statistically significant increase in IL-17A was observed in the plasma of FEP transition patients. In the multiple logistic regression model, higher IL-17A levels, combined with lower levels of MIP-3α and IFN-γ, were significantly associated with clinical stage transition. These findings emphasise the potential to develop a predictive model combining these three cytokines to better identify individuals at an increased risk of disease progression.

Integrating environmental and genetic factors into our multiple logistic regression analysis of cytokine biomarkers may improve model performance and provide a more comprehensive understanding of the factors influencing the transition of FEP.

One key limitation could be the small sample size, which might affect the statistical power of the analysis and increase the risk of a type II error. A larger sample size would enhance the ability to detect more subtle associations between cytokine levels and clinical stage transition.

Furthermore, an accuracy of 77.14% was achieved, demonstrating good performance in correctly classifying the samples. However, to improve diagnostic capacity, more variables could be added to the model, and cross-validation should be employed to prevent overfitting and ensure better generalisation.

Additionally, the predominantly male composition of both the T and nT groups introduces potential bias, which limits the generalisability of our findings. Although analysing only female participants would be highly valuable in further clarifying this interaction, it was not conducted due to the limited number of female participants in the cohort. Given the documented sex differences in immune responses and cytokine profiles in psychotic disorders [55, 56], future studies with a more balanced sample and larger female cohorts are necessary to better understand these dynamics and establish better comparisons between groups.

Moreover, the presence of multicollinearity, identified using the Variance Inflation Factor, especially between IL-17A and IFN-γ, can affect the accuracy and dependability of the results. This could have been avoided by increasing the sample size or removing one of these variables from the logistic regression model, although this approach might reduce the model’s sensitivity and specificity.

In conclusion, this study represents a real-world case with recruited individuals which were subsequently selected according to clinical criteria. Thus, it represents a seminal contribution to the field by demonstrating the potential of combined cytokine profiles as predictive biomarkers for clinical stage transition in FEP, laying the groundwork for improved early detection and intervention strategies. Further research is necessary to better understand the complex interactions between immune, genetic, and environmental factors influencing the progression of psychotic disorders. Exploring the specific roles of IL-17-producing cells and other immune modulators could be crucial for developing targeted therapeutic approaches for FEP and similar conditions.

## Conflicts of Interest

The authors declare no conflicts of interest.

## Author Contributions

MR, CE, VS performed data analysis; MR, CE wrote the manuscript; CS, MG performed data acquisition; MC, SM, MB, HC performed patient selection and sample collections; IB established and performed sample processing for biobanking; AP, AM, NM, BM supervised the work, secured funding and performed final proofing of the manuscript.

## Funding Statement

The authors would like to thank the financial support of the European Regional Development Fund (ERDF), through the COMPETE 2020 - Operational Programme for Competitiveness and Internationalisation and Portuguese national funds via FCT – Fundação para a Ciência e a Tecnologia, I.P., under projects: POCI-01-0145-FEDER-30943 (ref.: PTDC/MEC-PSQ/30943/2017), PTDC/MED-NEU/27946/2017, POCI-01-0145-FEDER-016428 (ref.: SAICTPAC/0010/2015), POCI-01-0145-FEDER-016795 (ref.: PTDC/NEU-SCC/7051/2014) – and UIDB/04539/2020, UIDP/04539/2020, LA/P/0058/2020 doi.org/10.54499/LA/P/0058/2020, and the National Mass Spectrometry Network (RNEM) [POCI-01-0145-FEDER-402-022125 Ref. ROTEIRO/0028/2013). MR was supported by a Ph.D. fellowship, https://doi.org/10.54499/2020.07749.BD and a research fellowship, IT137-25-049 (REPAIR - PL23-00001).CS was supported by a Ph.D. fellowship, SFRH/BD/88419/2012, co-financed by the European Social Fund (ESF) through the POCH- Programa Operacional do Capital Humano and national funds via FCT.

## Data Availability

All data produced in the present work are contained in the manuscript

## Notes

### Competing Interest Statement

The authors have declared no competing interest.

### Author Declarations

The authors assert that all procedures contributing to this work comply with the ethical standards of the relevant national and institutional committees on human experimentation and with the Helsinki Declaration of 1975, as revised in 2013. All procedures involving human subjects/patients were approved by the Ethics Committee of the Faculty of Medicine, University of Coimbra (CE-122/2015).

