## Supplementary Data for "IL-17A, IFN-γ, and MIP-3α Plasma Profiles Predict Clinical Stage Transition in First-Episode Psychosis"

**Full Institutional Mailing Addresses:**

CNC - Center for Neuroscience and Cell Biology – University of Coimbra

UC Biotech - Parque Tecnológico de Cantanhede, Núcleo 04, Lote 8

3060-197 Cantanhede – Portugal

**Affiliations:**

a. CNC - Center for Neuroscience and Cell Biology, University of Coimbra, Coimbra, Portugal
b. CIBB - Centre for Innovative Biomedicine and Biotechnology, University of Coimbra, Coimbra, Portugal
c. Faculty of Medicine of the University of Coimbra, University of Coimbra, Portugal
d. Psychiatry and Mental Health Department, Coimbra Local Health Unit, Portugal
e. CIBIT - Coimbra Institute for Biomedical Imaging and Translational Research, University of Coimbra, Portugal
f. Flow Cytometry Unit, Department of Clinical Pathology, Centro Hospitalar e Universitário de Coimbra, Coimbra, Portugal,
g. Coimbra Institute for Clinical and Biomedical Research, Faculty of Medicine, University of Coimbra, Coimbra, Portugal,
h. Instituto Politécnico de Coimbra, ESTESC-Coimbra Health School, Ciências Biomédicas Laboratoriais, Coimbra, Portugal

**Supplementary Figures**

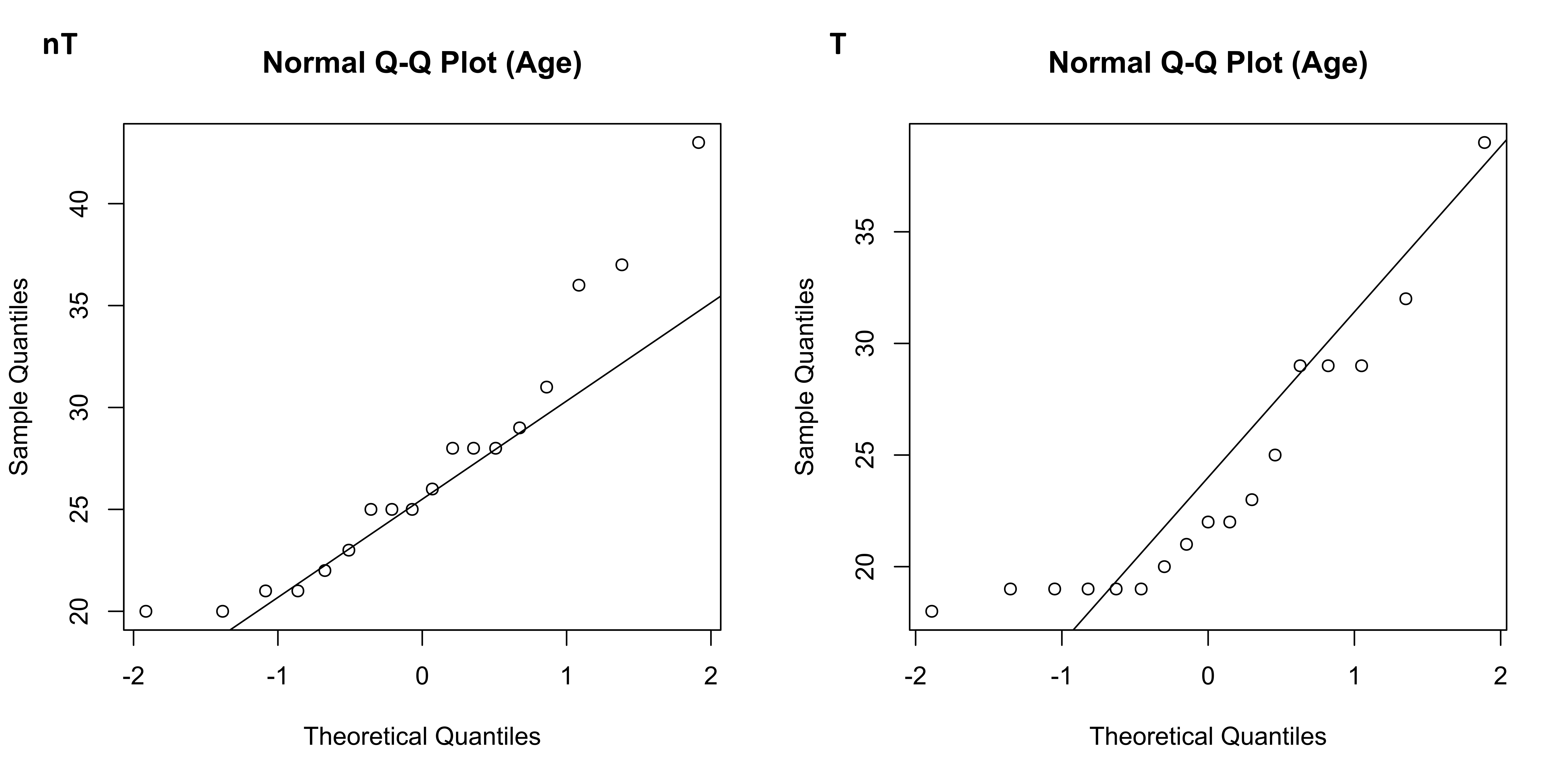
**Supplementary Figure 1.** Quantile–quantile plots for age of study participants.

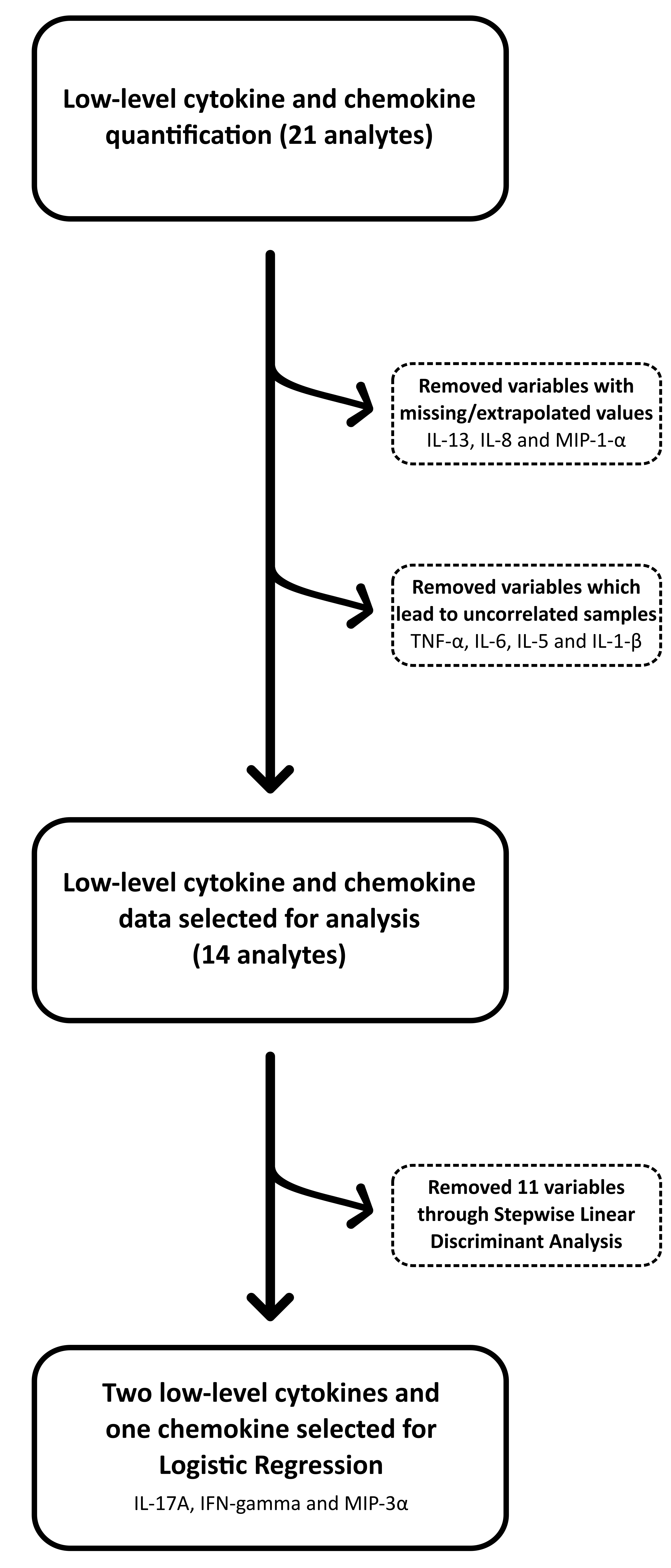

**Supplementary Figure 2.** Feature selection scheme.

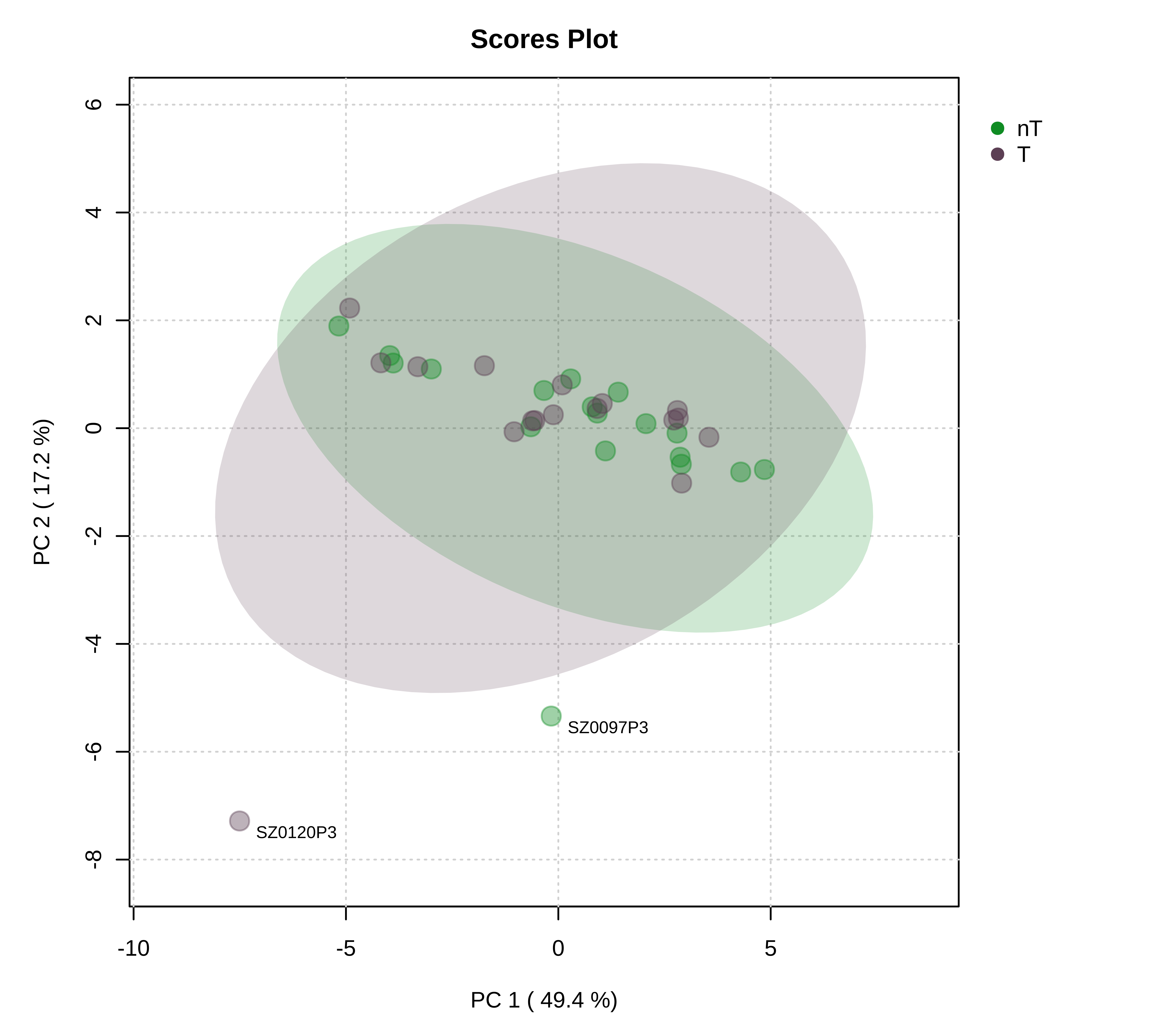

**Supplementary Figure 3.** Principal Component Analysis (PCA) using selected variables after removal of extrapolated/missing values.

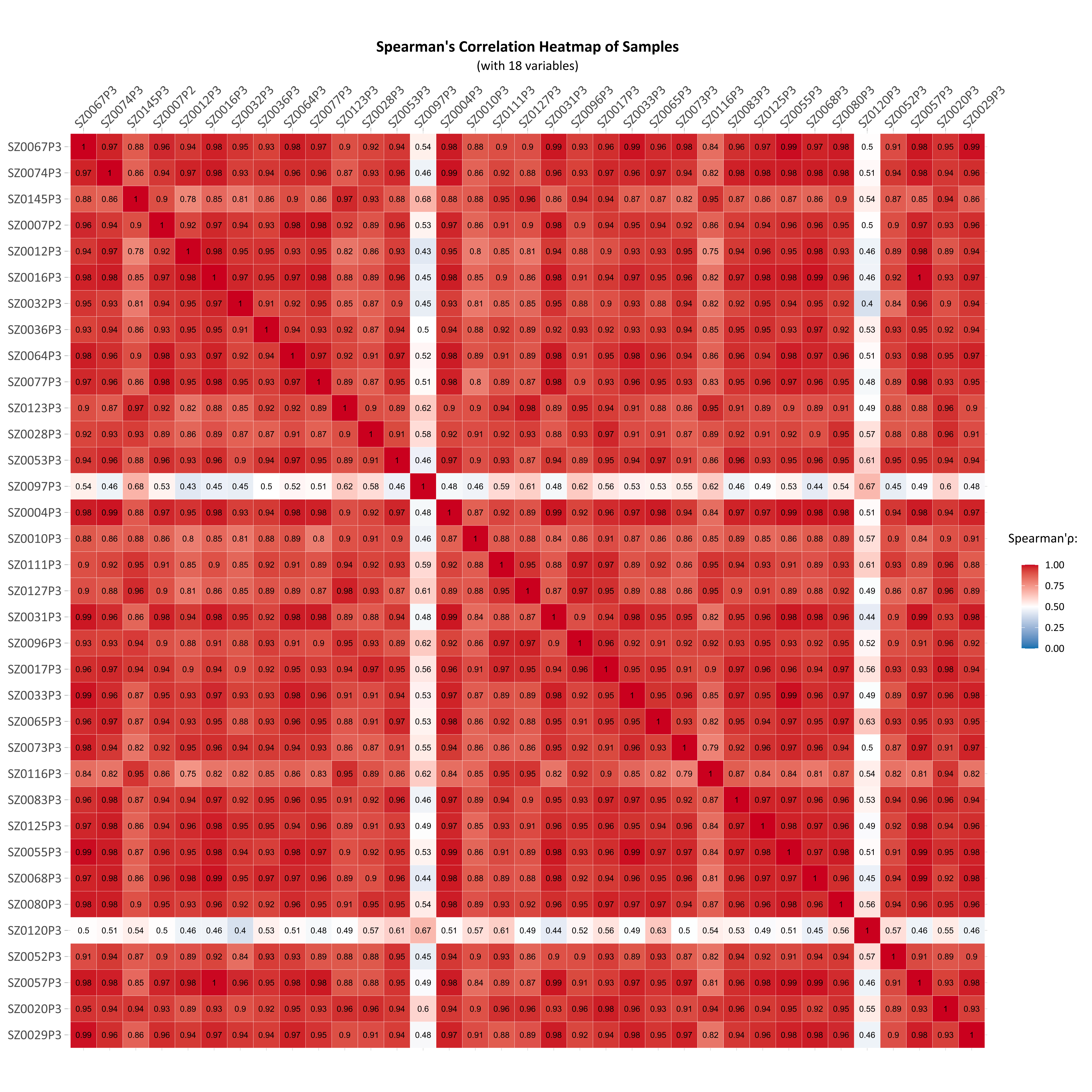

**Supplementary Figure 4.** Spearman’s correlation heatmap of samples using selected variables, after removal of extrapolated/missing values.

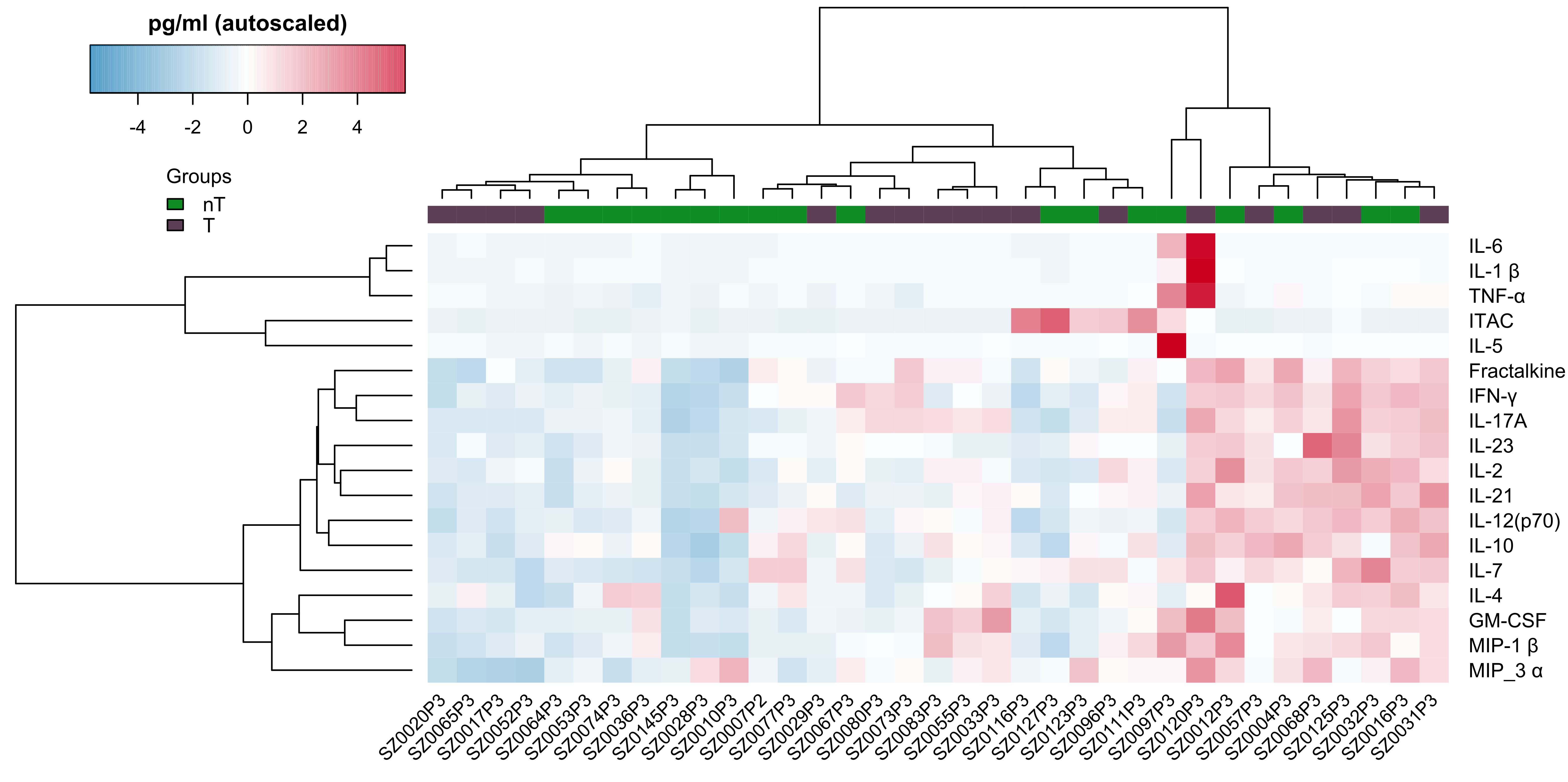

**Supplementary Figure 5.** Clustered heatmap of features with dendrograms after removal of extrapolated/missing values. Intensity values are expressed as standardised and mean-centred (autoscaled) values.

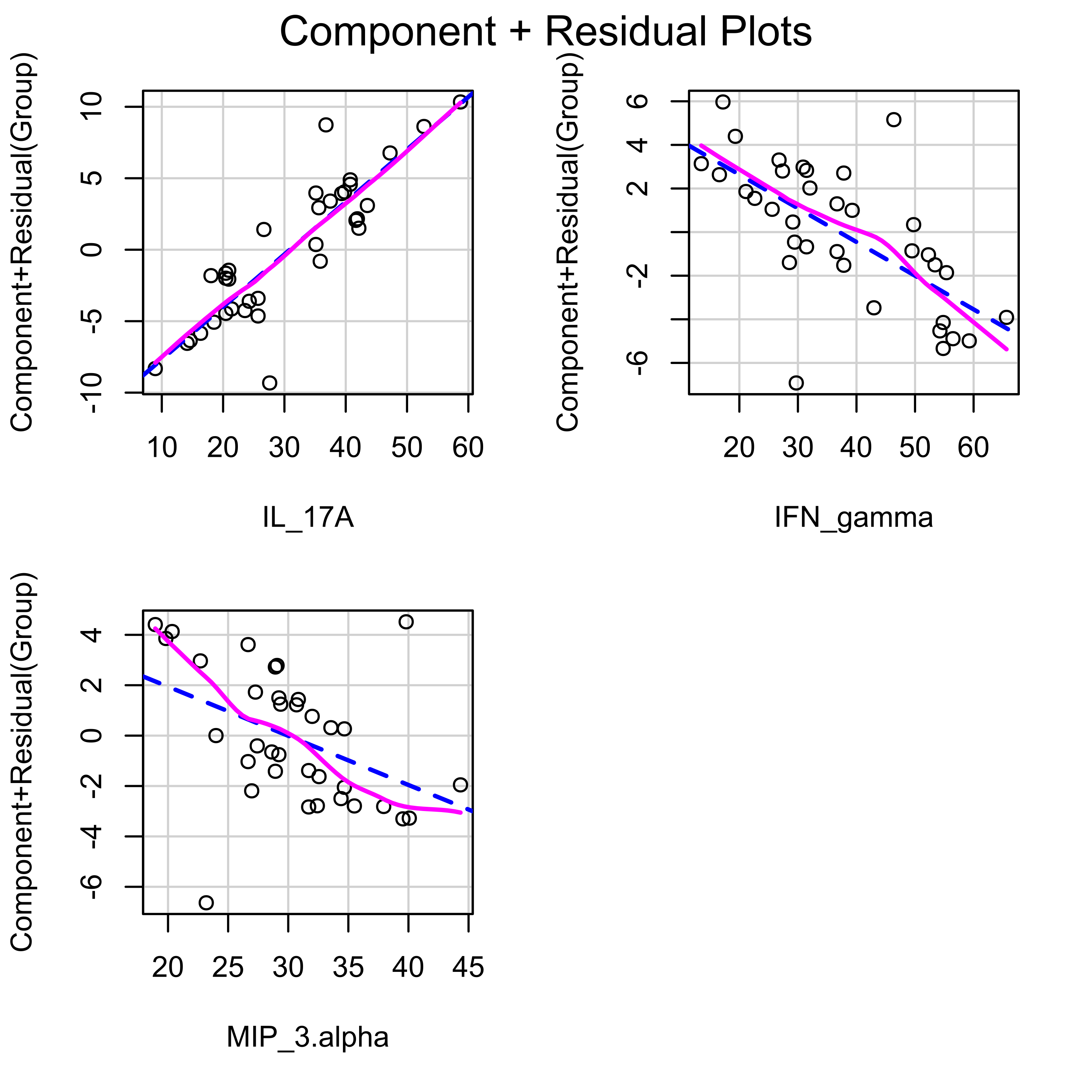

**Supplementary Figure 6.** Component-plus-residual plots of variables included in the logistic regression model.

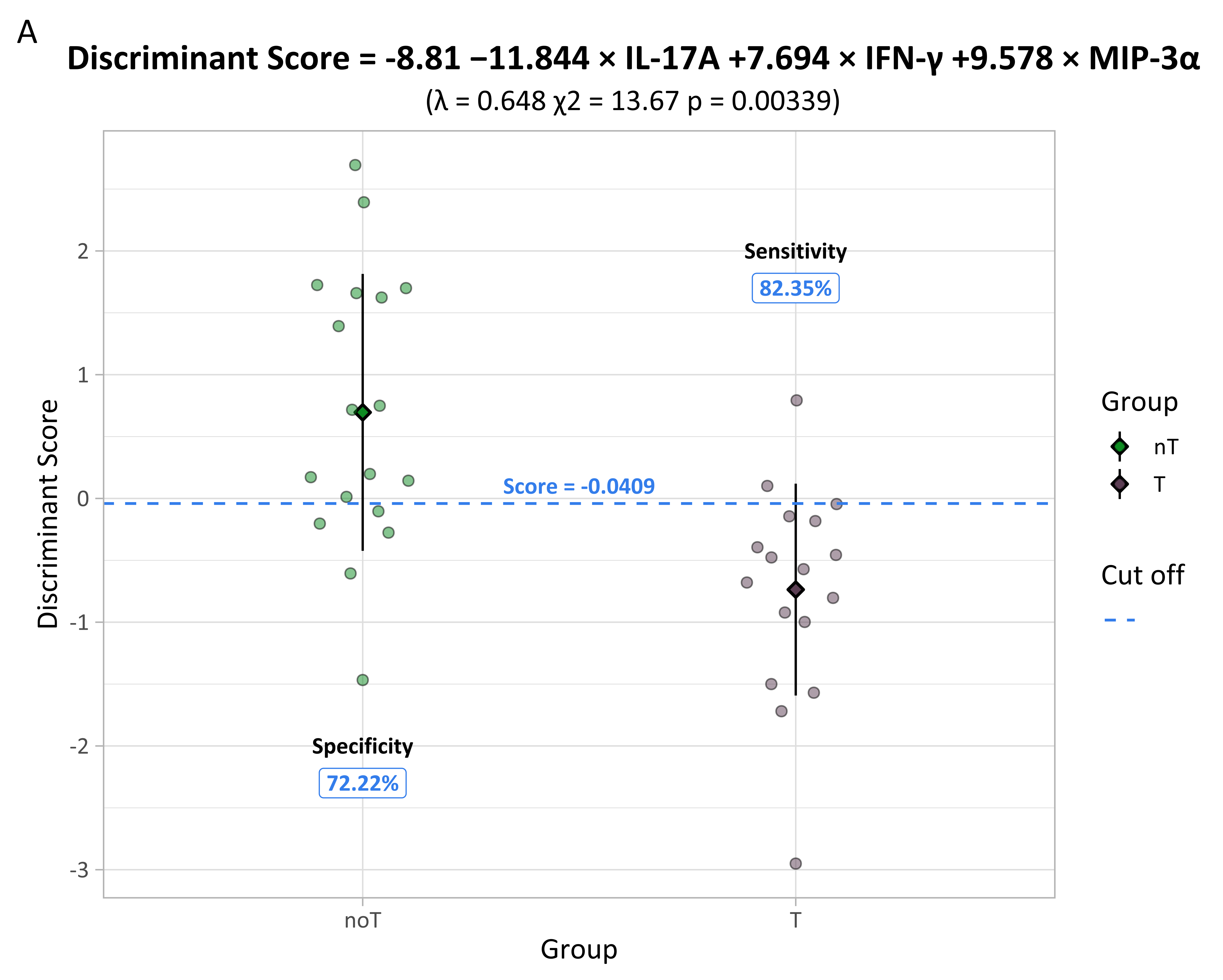

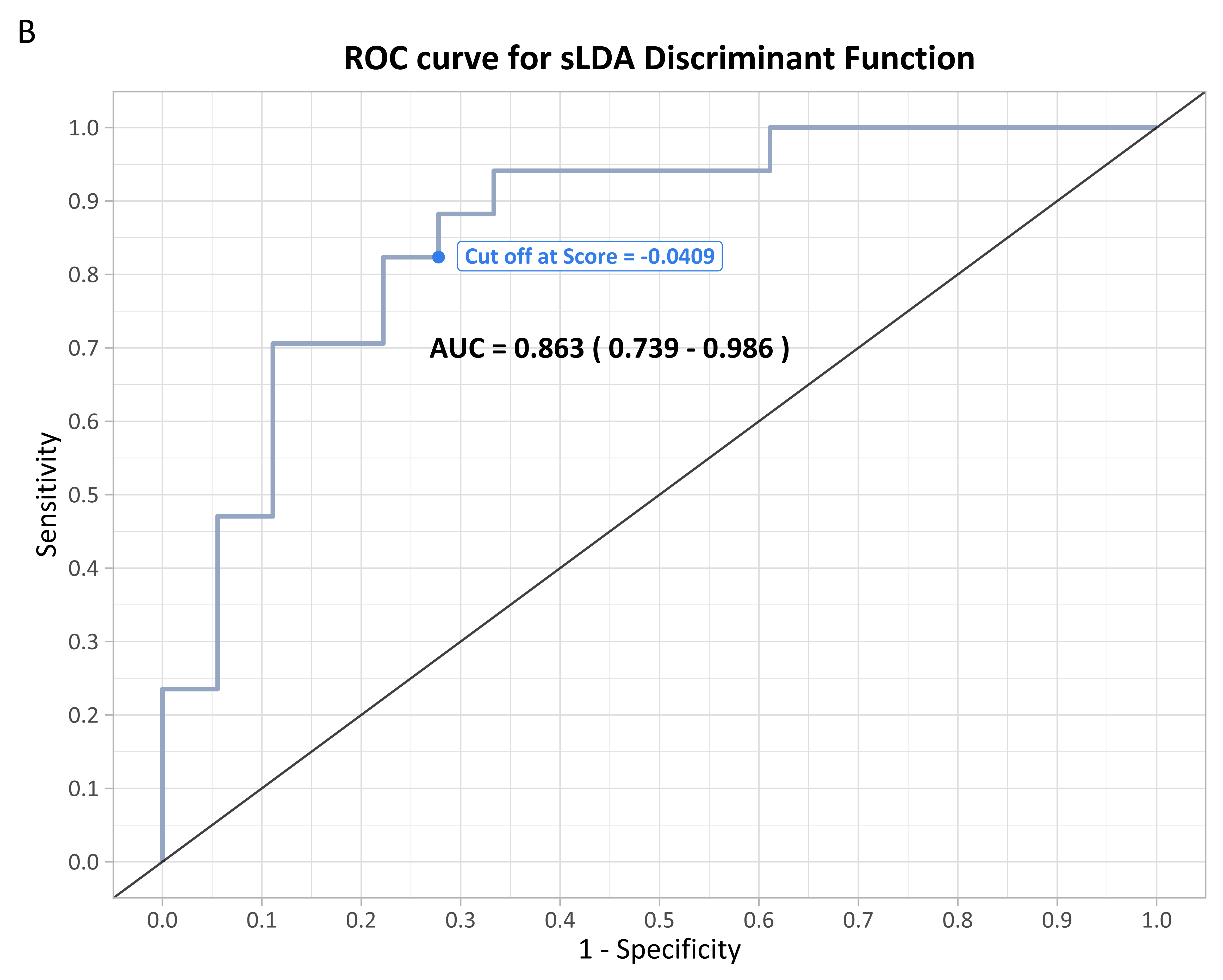

**Supplementary Figure 7.** Stepwise Linear Discriminant Analysis of pre-processed data. (A) Plot of discriminant function scores by group, with the classification cut-off indicated by a dashed blue line. Sensitivity and specificity values are shown for the chosen threshold. Diamonds represent group centroids, and vertical lines indicate the standard error. Log10-transformed data after pre-processing were used as input for the model. (B) Receiver Operating Characteristic curve for the discriminant function. The area under the curve and the corresponding 95% confidence interval are shown in bold black text. The classification cut-off score is marked by a blue dot.

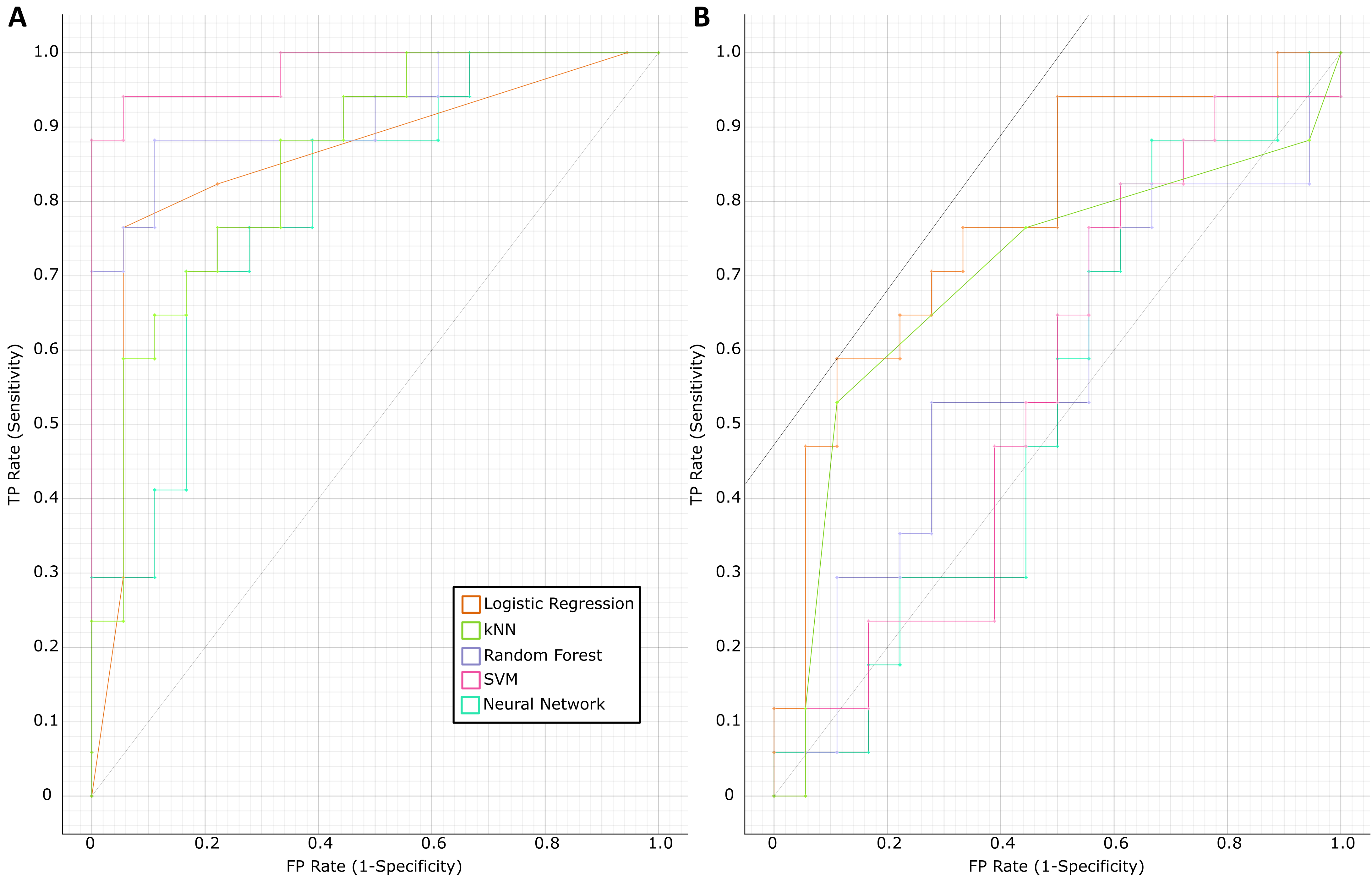

**Supplementary Figure 8.** Diagnostic Capacity of sLDA Selected Features Across Multiple Models. (A) Receiver Operating Characteristic curves of original models. (B) Receiver Operating Characteristic curves after Leave-One-Out Cross-Validation. TP – True Positive; FP – False Positive; kNN – K-Nearest Neighbours; SVM – Support Vector Machine.

**Supplementary Tables**

| **Supplementary Table 1. Univariate normality test on age of participants**  (Anderson-Darling test) | | | |
| --- | --- | --- | --- |
| **Group** | **Normality** | **Statistic** | **p-value** |
| **nT** | Normal | 0.638 | 0.081 |
| **T** | Not Normal | 1.036 | 0.007 |

| **Supplementary Table 7.**  **F Test to Compare Variances between study groups on normally distributed features.**  (Normally distributed features are those that are determined to be normal in both groups, as shown in Supplementary Table 5. ) | | |
| --- | --- | --- |
| **Analyte** | **Statistic** | **p-value** |
| **Fractalkine** | 1.299 | 0.606 |
| **IFN-γ** | 1.160 | 0.771 |
| **IL-10** | 1.156 | 0.777 |
| **MIP-3α** | 0.546 | 0.227 |
| **IL-12 (p70)** | 1.244 | 0.666 |
| **IL-17A** | 0.815 | 0.678 |
| **IL-4** | 3.594 | 0.014 |
| **IL-7** | 1.418 | 0.490 |
| **MIP-1β** | 2.305 | 0.102 |

| **Supplementary Table 9.**  **Multivariate normality test on selected features after data pre-processing.**  (Henze-Zirkler test) | | | |
| --- | --- | --- | --- |
| **Group** | **Normality** | **Statistic** | **p-value** |
| **nT** | Not Normal | 0.996 | 0.027 |
| **T** | Not Normal | 0.996 | 0.033 |

| **Supplementary Table 10.**  **Multivariate normality test on selected features after data pre-processing and log10 transformation.**  (Henze-Zirkler test) | | | |
| --- | --- | --- | --- |
| **Group** | **Normality** | **Statistic** | **p-value** |
| **nT** | Not Normal | 0.997 | 0.019 |
| **T** | Not Normal | 0.996 | 0.026 |

| **Supplementary Table 12. Logistic Regression Model Statistics** | | | | | | | | |
| --- | --- | --- | --- | --- | --- | --- | --- | --- |
|  | **Coefficients** | | |  | **Wald test** | | **Collinearity** | |
|  | **β** | **SE** | | **OR** | **z** | **p-value** | **Tolerance** | **VIF** |
| **Intercept** | 3.407 | | 2.345 | - | 1.452 | 0.146 | - | - |
| **IL-17A** | 0.332 | 0.124 | | 1.394 | 2.688 | 0.007 | 0.116 | 8.600 |
| **IFN-γ** | -0.176 | 0.086 | | 0.839 | -2.035 | 0.042 | 0.155 | 6.458 |
| **MIP-3α** | -0.232 | 0.107 | | 0.793 | -2.173 | 0.030 | 0.469 | 2.134 |

β – Coefficient; SE – Coefficient Standard Error; OR – Odds Ratio; z – Wald Test Statistic; VIF – Variance Inflation Factor.

| **Supplementary Table 13. Logistic Regression Model evaluation and goodness-of-fit** | | | |
| --- | --- | --- | --- |
|  | **χ^2^** | **DoF** | **p-value** |
| **Likelihood Ratio test** | 16.324 | 3 | >0.001 |
| **Hosmer and Lemeshow test** | 5.044 | 7 | 0.655 |

DoF – Degrees of Freedom

| **Supplementary Table 14. Logistic Regression Model with Box–Tidwell procedure** | | | | |
| --- | --- | --- | --- | --- |
|  | **Coefficients** | | **Wald test** | |
|  | **β** | **SE** | **z** | **p-value** |
| **Intercept** | 36.353 | 25.106 | 1.448 | 0.148 |
| **IL-17A** | -0.057 | 2.142 | -0.027 | 0.979 |
| **IFN-γ** | -0.077 | 1.673 | -0.046 | 0.963 |
| **MIP-3α** | -4.937 | 3.433 | -1.438 | 0.150 |
| **IL-17A * ln(IL-17A)** | 0.095 | 0.485 | 0.196 | 0.844 |
| **IFN-γ * ln(IFN-γ)** | -0.025 | 0.360 | -0.069 | 0.945 |
| **MIP-3α * ln(MIP-3α)** | 1.070 | 0.776 | 1.378 | 0.168 |

β – Coefficient; SE – Coefficient Standard Error; z – Wald Test Statistic.

| **Supplementary Table 15.**  **Classification Performance of sLDA Selected Features**  (Across Multiple Models) | | | | |
| --- | --- | --- | --- | --- |
| **Model** | **Original** | | **LOOCV** | |
|  | **AUC** | **CA** | **AUC** | **CA** |
| **Logistic Regression** | 0.853 | 0.771 | 0.778 | 0.686 |
| **kNN** | 0.855 | 0.857 | 0.698 | 0.714 |
| **Random Forest** | 0.977 | 0.914 | 0.569 | 0.600 |
| **SVM** | 0.918 | 0.800 | 0.549 | 0.600 |
| **Neural Network** | 0.801 | 0.714 | 0.533 | 0.486 |

kNN – K-Nearest Neighbours; SVM – Support Vector Machine; LOOCV – Leave-One-Out Cross-Validation; AUC – Area Under (Receiver Operating Characteristic) Curve; CA – Average Classification Accuracy over Classes.

| **Supplementary Table 17.** **Confusion matrices for sLDA determined discriminant function.** | | | | | |
| --- | --- | --- | --- | --- | --- |
|  | **Group** | **Original** | | **LOOCV** | |
|  |  | **nT** | **T** | **nT** | **T** |
| **Predicted Classification (n)** | **nT** | 13 | 3 | 13 | 5 |
|  | **T** | 5 | 14 | 5 | 12 |
| **Predicted Classification (%)** | **nT** | 72.2 | 17.6 | 72.2 | 29.4 |
|  | **T** | 27.8 | 82.4 | 27.8 | 70.6 |
| **Total Accuracy (%)** | | 77.1 | | 71.4 | |

LOOCV – Leave-One-Out Cross-Validation.

| **Supplementary Table 18.**  **Box's Test of Equality of Covariance Matrices for sLDA analysis.** | | | |
| --- | --- | --- | --- |
| **Box’s M** | | 9.489 | |
| **F** | **Approximation** | | 1.424 |
|  | **df1** | | 6 |
|  | **df2** | | 7813.106 |
|  | **p-value** | | 0.201 |
